# Cognitive Reserve Disrupts Cognitive Decline from White Matter Hyperintensities

**DOI:** 10.1101/2025.08.15.25331829

**Authors:** Arthur P. Hamilton, Kaiah N. Sotebeer, John G. Grundy, Katherine Chadwick, Cassandra Morrison, Mahsa Dadar, Ellen Bialystok, John A. E. Anderson, Alzheimer’s Disease Metabolomics Consortium

**Author notes:** Data used in preparation of this article were obtained from the Alzheimer’s Disease Neuroimaging Initiative (ADNI) database (adni.loni.usc.edu). As such, the investigators within the ADNI contributed to the design and implementation of ADNI and/or provided data but did not participate in analysis or writing of this report. A complete listing of ADNI investigators can be found at: http://adni.loni.usc.edu/wp-content/uploads/how_to_apply/ADNI_Acknowledgement_List.pdf. Contributed equally to the article (Arthur P. Hamilton, Kaiah N. Sotebeer).

## Abstract

Previous research examining the contribution of white matter hyperintensities (WMHs) to cognitive decline has focused on overall lesion burden. A new approach, afforded by the Lesion Quantification Toolkit (LQT), measures localized connectivity disruption from WMHs to better estimate their impact on cognition. This methodology shifts the focus from lesion volume to the level of network disruption between brain regions. In this novel study, we applied the LQT approach to healthy aging and linked the degree of disconnection of gray matter by WMHs to both cognitive impairment and resilience via cognitive reserve. Using three pre-existing MRI datasets of older adults (total N = 259), we used the LQT to examine localized disruptions to brain connectivity due to WMHs. We then used partial least-squares path modeling to examine the relationships between this disruption, cognitive performance, age, and cognitive reserve. The results support a link between connectivity disruption and reduced cognitive performance. An analysis of one of the three datasets, which included a detailed measure of cognitive reserve, showed a link between cognitive reserve and higher cognitive performance, suggesting cognitive reserve offsets the negative impact of WMHs.

**CRediT:** Conceptualization: JAEA, KNS, APH, JGG, CM; Data curation: APH, JAEA; Formal Analysis: APH, KC; Funding acquisition: JAEA; Investigation: APH; Methodology: APH, KNS; Project administration: APH, JAEA; Resources: MD, JAEA; Software: MD; Supervision: JAEA; Validation:; Visualization: APH; Writing – original draft: KNS, APH, JGG, JAEA; Writing – review & editing: APH, KNS, JGG, KC, CM, MD, EB, JAEA.

## 1. Introduction

The prevalence of health conditions associated with age-related cognitive decline, such as mild cognitive impairment and Alzheimer’s disease, continues to rise (Cova et al., 2017; Song et al., 2023). Such conditions typically lead to disability, reduced quality of life, and increased burden on health services. These conditions are suggested to occur as the culmination of a longer process of cognitive decline, which is linked with neurobiological mechanisms, including gray and white matter tissue loss as measured by volumetric decrease (Taki et al., 2011; Guttmann et al., 1998), cortical thinning (Fjell et al., 2014), reductions in fractional anisotropy (Grieve et al., 2007), and white matter hyperintensities (WMHs; Zhuang et al., 2018). Research has increasingly examined cognitive reserve, which is built by partaking in various cognitively engaging activities throughout the lifespan and is frequently associated with delayed cognitive decline. In the present study, the recently developed Lesion Quantification Toolkit (LQT; Griffis et al., 2021) was used to examine the impact of localized disruptions to connectivity due to WMHs on cognitive performance in older adults and how this impact may be altered by cognitive reserve.

Cognitive decline is a gradual reduction in several areas of cognitive ability, including working memory, attention, and episodic and associative memory. Most individuals begin the process of cognitive decline in their 30s, but there is significant individual variation (Salthouse, 2009). While cognitive decline is a normal part of healthy aging, it can lead to dementia, such as Alzheimer’s disease, and substantial functional impairment (Deary et al., 2009; Murman, 2015). Individuals with similar physiological indicators of cognitive decline often display different degrees of functioning, with the better performance of some individuals suggested to result from cognitive reserve (Tucker & Stern, 2011). Cognitive reserve is the resilience of cognitive function in the face of age-related brain pathology due to compensatory brain mechanisms (Pinter et al., 2015). Research on aging suggests that cognitive reserve may be bolstered by factors such as education (Meng & D’Arcy, 2012; Song et al., 2022), occupational attainment (Ghaffar et al., 2012), participation in leisure activities (Stern, 2012), and bilingualism (Bialystok, 2021). In general, cognitive reserve is helpful for offsetting cognitive decline.

A notable physiological predictor of cognitive decline is the development of WMHs—lesions that result from highly localized cerebral ischemia. WMHs are common in healthy aging, with over 90% of individuals developing WMHs by the age of 65 (d’Arbeloff et al., 2019). WMHs are caused by cerebral small vessel disease (also called leukoaraiosis; Black et al., 2009; Prins & Scheltens, 2015), the significant risk factors of which include hypertension, heart disease, and diabetes (Debette & Markus, 2010; Tamura & Araki, 2015). Small vessel disease is normal in healthy aging, but in high concentrations, it is associated with cognitive decline (Black et al., 2009). Meanwhile, WMHs interfere with neurotransmission along white matter tracts and thereby block communication between gray matter regions (Boot et al., 2020). WMHs can be detected through magnetic resonance imaging (MRI) using a fluid-attenuated inversion recovery sequence (Yoshita et al., 2005).

Prior research investigating the role of WMHs in aging has found an association between WMHs and an elevated risk of stroke (Giese et al., 2020). A meta-analysis including 31 prospective studies on WMHs and dementia (Hu et al., 2021) found that WMHs were a significant risk factor for dementia (relative risk [RR]: 1.12), including Alzheimer’s disease (RR: 1.25) and vascular dementia (RR: 1.73). Furthermore, Smith et al. (2011) found that increased WMH load is associated with reduced executive function and episodic memory performance in older adults after accounting for age and education. Similarly, Kamal et al. (2023) found that higher WMH load is associated with lower global cognition, executive function, and memory scores in cognitively normal older adults, and that older adults with mild cognitive impairment or Alzheimer’s disease had higher WMH load than cognitively normal adults.

The findings of two meta-analyses support an overall negative effect of WMHs on cognitive ability. First, a meta-analysis of 17 MRI studies on WMHs and cognition found that for cognitively normal older adults, the presence of WMHs yielded a significant effect on cognition as measured through validated neuropsychological tests such as the Mini-Mental State Examination (MMSE; Kloppenborg et al., 2014). WMHs were also found to be associated with general memory decline (*z* = −0.07) and a greater decline in executive functioning (*z* = −0.32), including attention and processing speed. Second, a meta-analysis of prospective studies on WMHs as a risk factor identified five studies on cognitive impairment and WMHs that found that the presence of WMHs at baseline increased the RR of cognitive impairment (RR: 1.27; Hu et al., 2021).

However, the presence of WMHs does not always indicate cognitive impairment or dementia symptoms, and some individuals experience more rapid decline due to WMHs than others (Lampe et al., 2019). Indeed, there is some variability in the literature surrounding the role of WMH volume in general cognitive decline (Coenen et al., 2023; Kim et al., 2020). Furthermore, one study investigating the association between WMH density and cognition showed only selective age-related declines in executive functioning and no decline in working memory performance (Boutzoukas et al., 2021).

Recently, researchers have begun to examine which regions most adversely impact cognitive ability when disconnected due to WMHs. Assessing the localization of WMHs as an alternative to whole-brain analyses may provide more detailed insights into the role of WMHs in cognitive decline. In addition, a more thorough investigation into the localization of WMHs may explain the variations between studies regarding the extent to which WMHs contribute to cognitive decline. More recent literature on WMHs and cognitive decline frequently focuses on the anterior cingulate cortex and the frontal lobes due to their association with higher-order executive functions (Wang et al., 2022; Anor et al., 2021). Unlike traditional whole-brain approaches, these studies help inform how WMHs in specific regions are more likely to contribute to cognitive decline.

The LQT (Griffis et al., 2021) is a software package developed to identify disconnection caused by WMHs at the levels of individual white matter tracts and gray matter regions individual parcels (i.e., brain regions as defined by a chosen atlas). The LQT, which was originally created to measure structural connectivity disruption due to WMHs in ischemic stroke patients (Griffis et al., 2021), provides estimates of disruption to each white matter tract and communication to each gray matter region. Griffis et al. (2021) successfully related these connectivity disruption measures to cognitive performance in older adults in sub-acute stroke (i.e., the period beginning two weeks after a stroke) and found that the presence of stroke lesions intersecting specific white matter pathways was predictive of specific cognitive deficits. For example, language abilities were reduced in patients with WMHs in the left arcuate fasciculus and the left inferior fronto-occipital fasciculus.

However, there is a lack of research addressing whether differences in the LQT’s connectivity disruption measures predict reduced cognitive performance in cognitively normal older adults. Therefore, the aim of the present study was to investigate the impact of WMHs on cognitive performance in a sample of cognitively normal older adults using the LQT to expand upon prior research on WMHs and cognitive decline by examining the predictive value of detailed parcel-level measures of disrupted connectivity. Our goals were to 1) assess the potential value of the LQT for studies on cognitively normal older adults, 2) examine the relationship between cognitive performance and the LQT’s measure of disrupted gray matter connectivity, and 3) examine how cognitive reserve affects this relationship.

## 2. Methods

### 2.1 Datasets

Data were obtained from three pre-existing sources: 1) a dataset collected by the Alzheimer’s Disease Neuroimaging Initiative (ADNI), 2) an unpublished portion of a dataset collected by E.B.’s lab for other purposes at York University, and 3) an open-access dataset from Cornell University (Spreng et al., 2022). All data used for this study were cross-sectional. The term “combined” dataset refers to all participants from the three individual datasets together.

Data used in the preparation of this article were obtained from the Alzheimer’s Disease Neuroimaging Initiative (ADNI) database (adni.loni.usc.edu). The ADNI was launched in 2003 as a public-private partnership, led by Principal Investigator Michael W. Weiner, MD. The primary goal of ADNI has been to test whether serial MRI, positron emission tomography (PET), other biological markers, and clinical and neuropsychological assessment can be combined to measure the progression of mild cognitive impairment (MCI) and early Alzheimer’s disease (AD).

For the datasets, from ADNI and Cornell, only subsets were used. The ADNI data used in this study were from ADNI-3, which was collected from 2016–2021. Only participants from ADNI-3’s second time point (2017) have been included, as this was the first time point including data from both new participants and participants continuing from earlier phases of the study. Furthermore, only ADNI participants assessed as cognitively normal (i.e., without a diagnosis of mild cognitive impairment or dementia) for that time point have been included (for full details on how the ADNI participants were screened at the download stage, see the Supplementary Material). The Cornell dataset is divided into younger and older groups; only the older group was used in this study.

For all datasets, the following demographic and behavioral measures were included: age, sex, education in years, and total score on the Mini-Mental State Examination (MMSE; Folstein et al., 1975). For each dataset, a set of cognitive task scores not shared with the other datasets was also included (see Table 1); this was employed instead of the MMSE for analyses of the individual datasets but not for combined analysis. For the ADNI and Cornell datasets, only years of education were available to measure cognitive reserve, while the York dataset includes a more detailed cognitive reserve measure, the Cognitive Reserve Index Questionnaire (CRIq; Nucci et al., 2012).

**Table 1.** Description of Cognitive Reserve Measures and Cognitive Tasks by Dataset.

|  | ADNI | York | Cornell | Combined |
| --- | --- | --- | --- | --- |
| Cognitive Ability Measures | Composite scores for<br>• memory<br>• executive functioning<br>• language<br>• visuospatial reasoning | • Shipley-2 (Shipley et al., 2009)<br>• Trail-Making Test (Army Individual Test Battery, 1944)<br>• Color–Word Interference Test (Stroop, 1935)<br>• Verbal Fluency Test (see Lezak et al., 2004 for review) | Composite scores for<br>• semantic memory<br>• episodic memory<br>• executive function | • Mini-Mental State Examination (MMSE; Folstein et al., 1975) |
| Cognitive Reserve Measures | • Education (years) | • Cognitive Reserve Index Questionnaire (Nucci et al., 2012) | • Education (years) | • Education (years) |

### 2.2 Participants

Participants were deemed ineligible for two reasons. Firstly, in the ADNI dataset, they were removed if they did not have a diagnosis of “Cognitively Normal” (i.e., they had a diagnosis of mild cognitive impairment, a diagnosis of dementia, or no diagnosis listed) or if they were removed if they were from the “Young” group of participants in the Cornell dataset. The number of participants excluded at each stage of data analysis is shown in Figure 1, and descriptive statistics of the final sample are shown in Figure 2.

**Figure 1.**
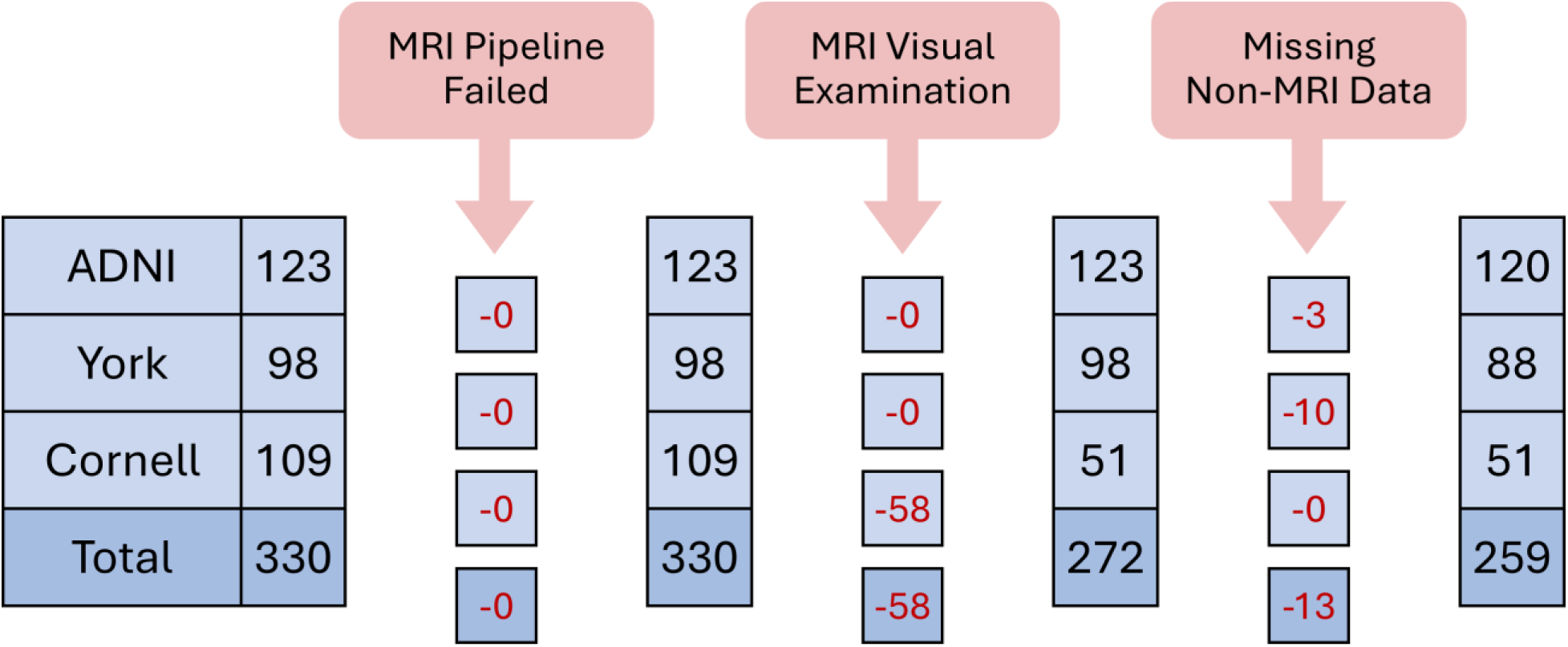
Exclusion of Participants by Data Analysis Stage by Dataset. *Note.* Number of participants excluded and remaining at each stage of data analysis. The leftmost column indicates the number of participants whose MRI scans were analyzed. The columns with red text indicate the number of participants who were removed at a given stage, while the columns with black text indicate the number of participants still included. Note that 88 participants from the York dataset were included in the combined dataset (as shown in the figure above) after excluding those with missing data necessary for the combined dataset; one additional participant was excluded from the York only dataset based on missing data, leaving a final sample of 87 participants for analyses.

**Figure 2.**
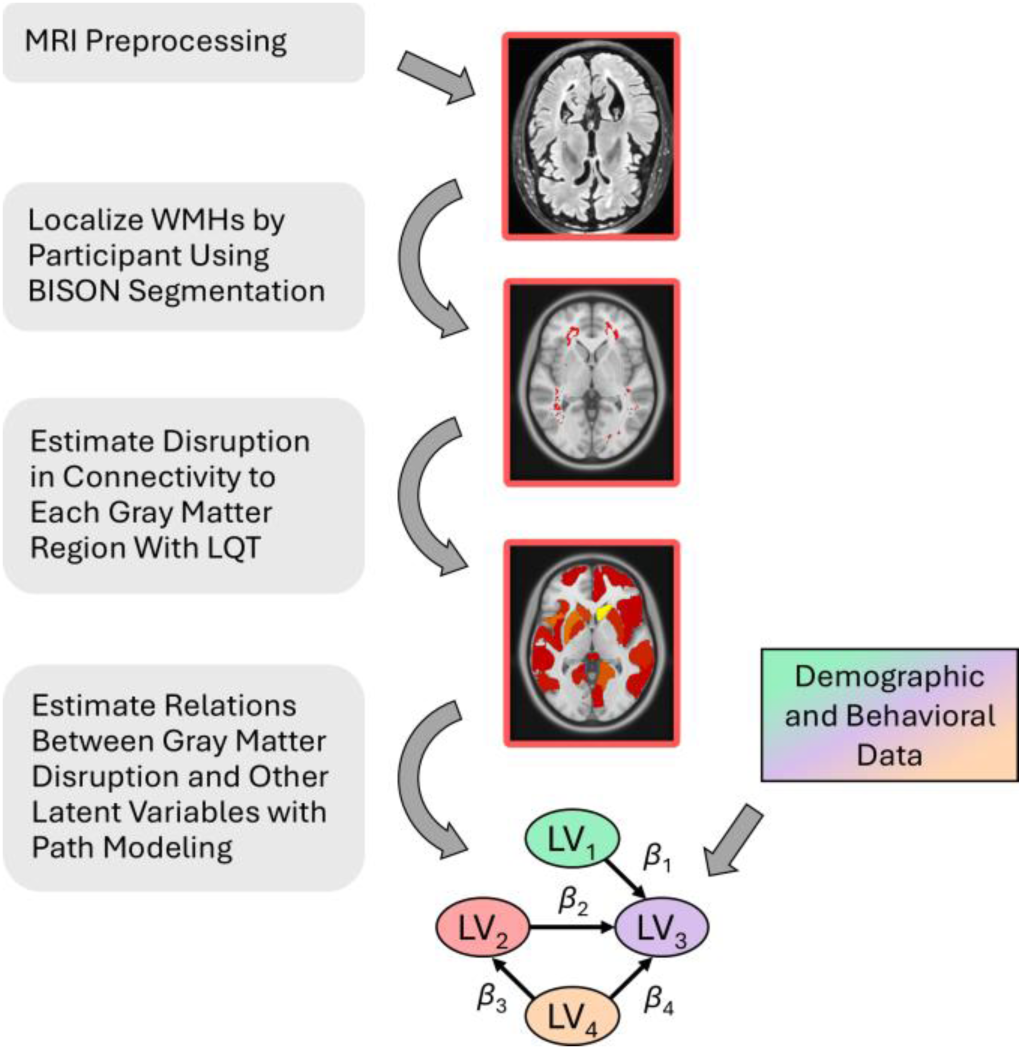
Summary of Key Data Processing Steps. *Note*. The box-and-arrow plot at the bottom is a partial least-squares path model (PLS-PM); LV indicates a latent variable; **β** indicates a coefficient for a path between two LVs. LV_1_ = cognitive reserve; LV_2_ = gray matter disruption; LV_3_ = cognitive ability; LV_4_ = age. Regional gray matter disruption scores are outputs from the LQT, while the other LVs are demographic and behavioral data and are only introduced at this stage of the analysis.

### 2.3 MRI Processing

The data were preprocessed using a previously validated pipeline that is publicly available on GitHub (https://github.com/VANDAlab/Preprocessing_Pipeline). This included denoising, intensity normalization, and image co-registration to the T1-weighted MRI and WMH segmentation using BISON segmentation (Dadar & Collins, 2021). After preprocessing, the segmented images were visually assessed for quality assurance. The segmentations were non-linearly registered to the MNI ICBM 152 template space. Using the MNI ICBM 152 gray matter and CSF atlas, voxels in gray matter and cerebrospinal fluid areas (i.e., ventricles) were excluded to remove partial volume effects.

The lesion maps were further processed by the LQT (Griffis et al., 2021) to estimate the level of disruption to connectivity caused by WMHs for individual white matter tracts and gray matter regions. In this study, the brain’s white matter was parcellated into 70 tracts based on the atlas of Yeh et al. (2018), and the brain’s gray matter was parcellated into 135 regions based on the Schaefer atlas (Schaefer et al., 2018). Disruption to a white matter tract was calculated as the percent disconnection severity matrix relative to the atlas structural connectivity matrix (Griffis et al., 2021). Gray matter disruption to a voxel was calculated as the proportion of streamlines intersecting it that had WMHs, and disruption to a gray matter region was calculated as the average disruption across voxels in the region.

### 2.4 Statistical Analysis

Analyses were not preregistered but data used for the statistical analyses and outputs from the analyses are available on OSF (https://osf.io/px7dy/?view_only=af90da9374ab44ea8fc47118817bc824). Statistical analyses were conducted in R on the ADNI, York, and Cornell datasets, and the combined dataset. The combined dataset was harmonized to remove dataset-specific sources of variance from the gray matter disruption scores using NeuroComBat (Fortin et al., 2018), a neuroimaging implementation of the ComBat harmonization method (Johnson et al., 2007). Variance associated with differences in the distribution of sex between datasets was preserved during harmonization.

The modeling was completed using partial least-squares path modeling (PLS-PM; Wold, 1975), which combines partial least-squares regression and structural equation modeling. PLS-PM was implemented using the plspm package in R (Sanchez, 2013). PLS-PM begins with a structural model of directional causal relationships (i.e., paths) between latent variables, which is defined a priori by the researcher. The strengths of the relationships in the structural model are then estimated using a measurement model, which relates the latent variables to observed variables for which empirical data is available. For all datasets, the structural model was the same and included four latent variables: cognitive performance, gray matter disruption, age, and cognitive reserve. For all datasets, these variables were connected by the same four paths: gray matter disruption to cognitive performance, age to gray matter disruption, age to cognitive performance, and cognitive reserve to cognitive performance.

The measurement model differed between datasets. For all datasets, overall gray matter disruption scores were derived from the disruption scores for the 135 gray matter regions. For the combined dataset, cognitive performance was represented by participant total scores on the MMSE (Folstein et al., 1975), the only cognitive assessment common to all three datasets. For the individual datasets, cognitive performance was derived from scores on a greater range of cognitive assessments available for each dataset (i.e.,Verbal Fluency Test). For the combined, ADNI, and Cornell datasets, years of education was the proxy for cognitive reserve, while for the York dataset, cognitive reserve was derived from the CRIq subscale scores (Nucci et al., 2012).

The resulting PLS-PM model for each dataset included coefficients for the strength of each of the four paths between variables specified previously. In addition, each model was bootstrapped 500 times, and a 95% bootstrap confidence interval was constructed. Paths were regarded as statistically reliable if their 95% bootstrap confidence interval did not include zero.

## 3. Results

The path from gray matter disruption to cognitive ability was statistically reliable in the combined dataset and the ADNI dataset, with greater gray matter disruption associated with lower cognitive ability. The strengths of the paths connecting variables in the PLS-PM models for each dataset are shown in Table 3. Visualizations of the PLS-PM models for the combined dataset and the York dataset are shown in Figures 3A and 3B, respectively; we focus on the York dataset because it has the most comprehensive representation of cognitive reserve. Visualizations of the ADNI and Cornell datasets can be found in the supplementary material in Figures S1A and S1B, respectively. In all datasets (including the combined dataset), the path from age to gray matter disruption was statistically reliable, with older age associated with greater gray matter disruption.

**Figure 3A.**
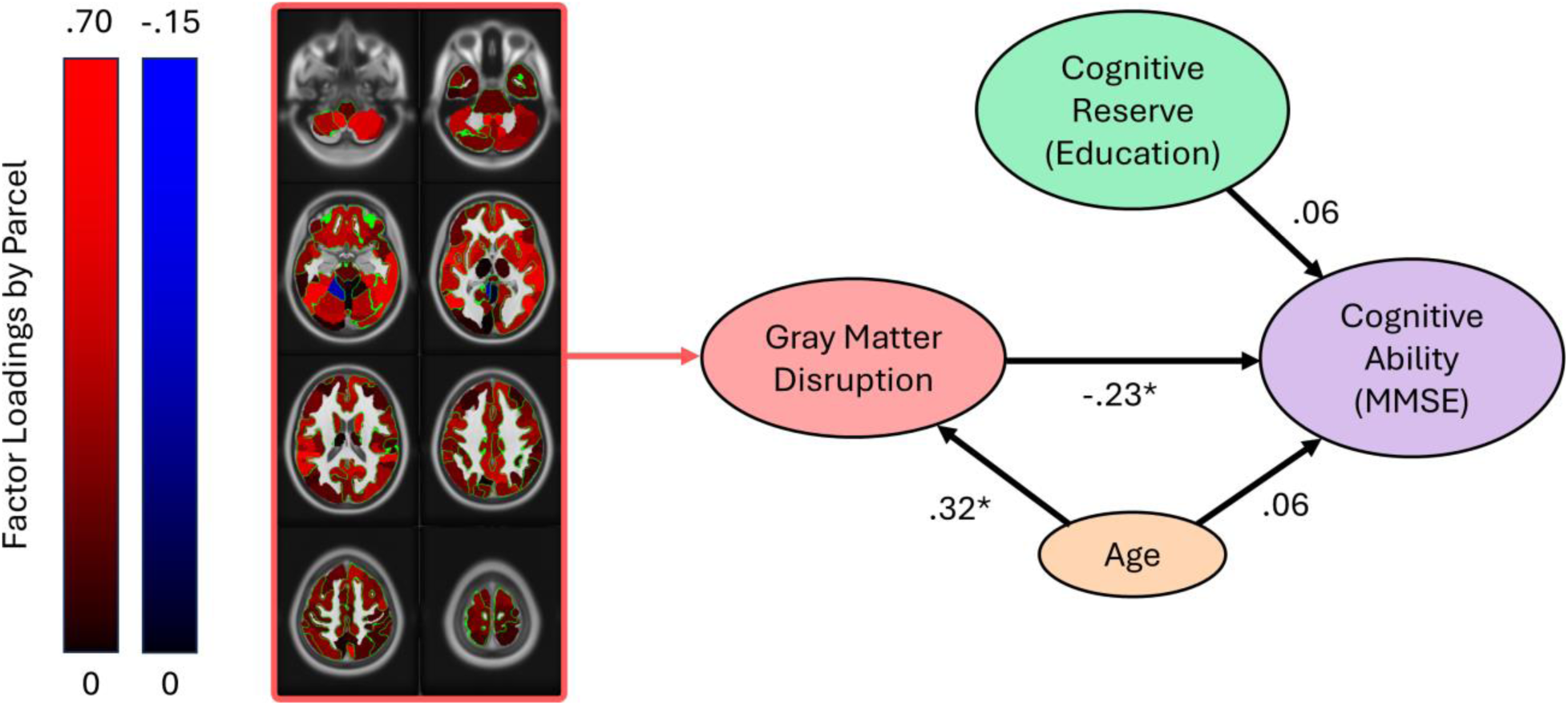
PLS-PM Model for the Combined Dataset. *Note.* This PLS-PM model is based on the combined dataset (i.e., including the ADNI, York, and Cornell datasets) after NeuroComBat harmonization (*N* = 259). It includes only behavioral measures shared between all three datasets. The black arrows represent the directional causal paths between variables specified a priori from past literature for the path model. The coefficients beside the arrows represent the strength of the causal links determined empirically from the data using PLS-PM. Asterisks next to coefficients indicate statistically reliable paths (i.e., the bootstrapped 95% confidence intervals for the coefficients did not include zero). Gray matter disruption indicates the extent to which communication to gray matter regions in the brain is affected by WMHs in white matter tracts connecting to them. The loadings of disruption scores for individual gray matter regions onto the model’s overall gray matter disruption factor are shown on the left. Positive loadings are shown in red, and negative loadings are shown in blue; brighter shades represent greater absolute values.

**Figure 3B.**
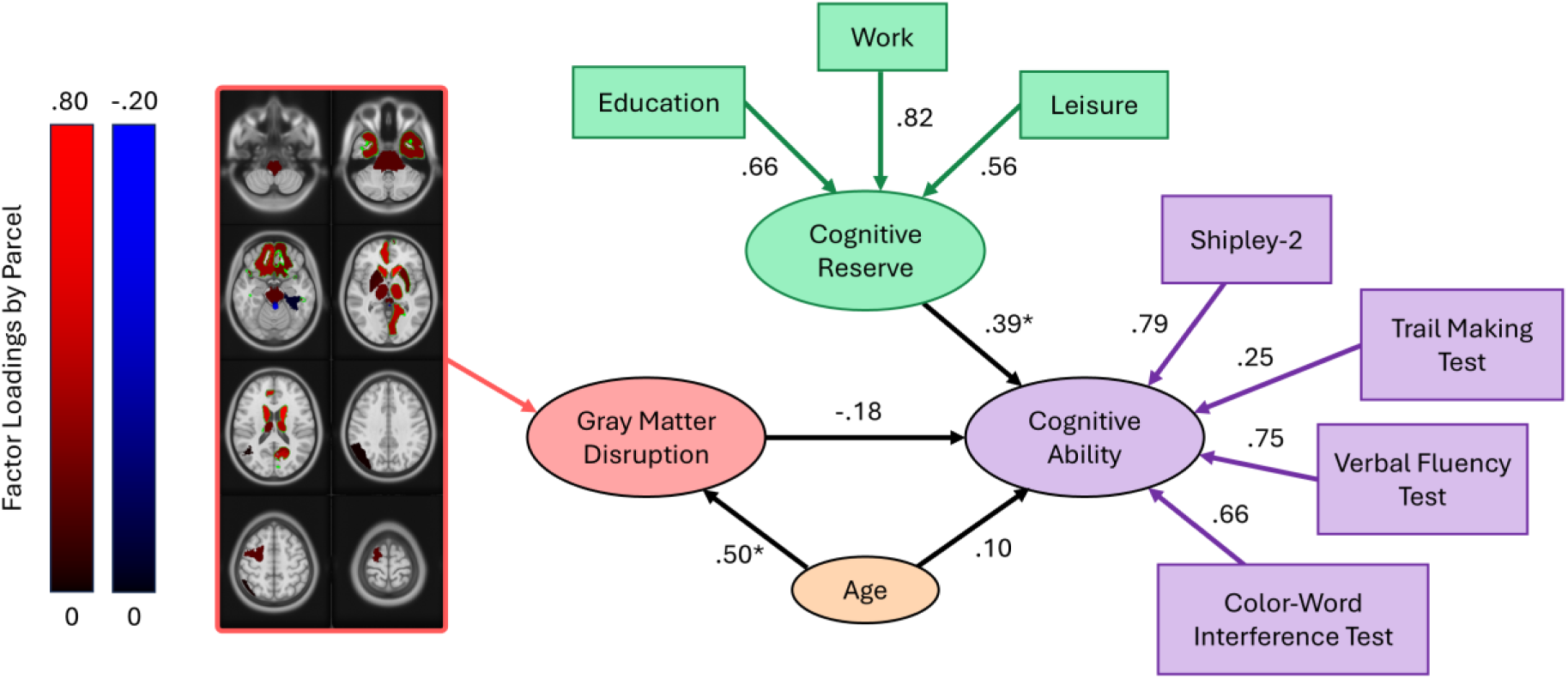
PLS-PM Model for the York Dataset. *Note.* This PLS-PM model is based on the York dataset only (*N* = 87). The black arrows represent the directional causal paths between variables specified a priori from past literature for the path model. The coefficients beside the arrows represent the strength of the causal links determined empirically from the data using PLS-PM. Asterisks next to coefficients indicate statistically reliable paths (i.e., the bootstrapped 95% confidence intervals for the coefficients did not include zero). Gray matter disruption indicates the extent to which communication to gray matter regions in the brain is affected by WMHs in white matter tracts connecting to them. The loadings of disruption scores for individual gray matter regions onto the overall gray matter disruption factor are shown on the left. Positive loadings are shown in red, and negative loadings are shown in blue; brighter shades represent greater absolute values.

**Table 2.** Sample Characteristics by Dataset.

| Variable | ADNI | York | Cornell | Combined |
| --- | --- | --- | --- | --- |
| *N* | 120 | 88 | 51 | 259 |
| Age | 70.16 (6.68) | 74.99 (4.07) | 69.45 (7.25) | 71.66 (6.49) |
| Sex | 68F, 52M | 62F, 26M | 35F, 16M | 165F, 94M |
| Education (years) | 16.79 (2.21) | 16.05 (2.64) | 16.37 (2.63) | 16.46 (2.46) |
| MMSE | 29.06 (1.16) | 29.28 (0.98) | 28.57 (1.40) | 29.04 (1.18) |

*Note.* For age, education, and MMSE, the values indicate $M$ ( $SD$ ).

**Table 3.** PLS-PM Path Coefficients by Dataset.

| Path | <u>Age to GMD</u> | <u>Age to Cog.</u> | <u>CR to Cog.</u> | <u>GMD to Cog.</u> |
| --- | --- | --- | --- | --- |
| Dataset | *M (95% CI)* | *M (95% CI)* | *M (95% CI)* | *M (95% CI)* |
| Combined | **0.32 (0.25, 0.50)** | 0.06 (-0.03, 0.25) | 0.06 (-0.05, 0.17) | **-0.23 (-0.45, -0.19)** |
| ADNI | **0.43 (0.34, 0.58)** | **-0.34 (-0.50, -0.17)** | **0.22 (0.01, 0.39)** | **-0.16 (-0.36, -0.04)** |
| York | **0.50 (0.38, 0.66)** | 0.10 (-0.22, 0.33) | **0.39 (0.21, 0.54)** | -0.18 (-0.54, 0.34) |
| Cornell | **0.79 (0.67, 0.89)** | 0.27 (-0.67, 0.74) | **0.33 (0.08, 0.61)** | -0.07 (-0.85, 0.60) |

*Note.* GMD = mean gray matter disruption. Cog. = cognitive ability. CR = cognitive reserve. Results marked in bold are statistically reliable based on their 95% bootstrap confidence intervals.

The path from cognitive reserve to cognitive ability was statistically reliable for all datasets, with the exception of the combined dataset. Greater cognitive reserve was associated with greater cognitive ability. The York dataset, which had the most detailed measure of cognitive reserve, showed the strongest effect.

Further details on the PLS-PM results can be found in the Supplementary Material in Table S1, Table S2, and Figure S2. Table S1 shows results for PLS regression to predict cognitive ability using all variables, for the four datasets. Table S2 shows indicators of unidimensionality for the latent variable for gray matter disruption, which describe the extent to which disruption scores for the 135 gray matter regions can be interpreted meaningfully as a single indicator. The values for Cronbach’s alpha (Cronbach, 1951) and Dillon-Goldstein’s rho (Dillon & Goldstein, 1984) for all datasets were above 0.70, a threshold recommended for PLS-PM analysis (Sanchez, 2013). However, the Dillon-Goldstein’s rho was 0.69 for the Cornell dataset, below this threshold, suggesting results relating to gray matter disruption for this dataset should be interpreted with caution. Figures S2 provides further detail on the combined dataset by showing PLS-PM models run with the same variables as the combined model (only MMSE for cognitive ability, only education for cognitive reserve) but only the participants from one dataset in each pane (ADNI for Figure S2A, York for Figure S2B, and Cornell for Figure S2C).

## 4. Discussion

We analyzed three pre-existing structural MRI datasets of cognitively normal older adults to examine the effect of connectivity disruption from WMHs on cognitive ability and how that relationship was affected by cognitive reserve. Connectivity disruption was measured with the LQT, a recent software package that can estimate the extent to which communication to gray matter regions is disrupted by WMHs in adjoining tracts. We created PLS-PM models to examine the effects of gray matter disruption and cognitive reserve on cognitive ability while also accounting for the effects of age on gray matter disruption and cognitive ability. The datasets were assessed individually and as a combined dataset. The PLS-PM model for the combined dataset showed that gray matter disruption was a statistically reliable predictor of lower cognitive ability. The York dataset contained a detailed measure of cognitive reserve, which was a statistically reliable predictor of higher cognitive ability.

### 4.1 Implications

Across all datasets, general cognitive functioning, measured with the MMSE, was negatively associated with WMH, and cognitive reserve had a minimal impact. Notably, in the York dataset, cognitive ability was primarily associated with verbal fluency, inhibition, visuospatial reasoning, and fluid intelligence; the trail making test, which assesses mental flexibility, attention, and speed, was less important to this construct.

Gray matter disruption scores obtained from analysis of WMHs in the combined dataset were a statistically reliable predictor of cognitive ability in older adults, even with the role of age and education accounted for. Nearly all gray matter regions loaded positively on the overall gray matter disruption factor, with bilateral subcortical regions loading the most strongly.

A model based on the York dataset, which included responses to the CRIq (Nucci et al., 2012), a detailed measure of cognitive reserve, showed a reliable positive effect of cognitive reserve on cognitive ability. This finding corroborates existing research supporting a protective effect of participating in cognitively engaging activities throughout the lifespan. In addition, this finding suggests that despite the importance of cognitive reserve, single-dimensional measures, such as years of education, may not be sufficient to assess it effectively. All three CRIq subscales (life history of education, work, and leisure) had high factor loadings on the overall cognitive reserve factor for the York model, but work had the highest loading. This may be because participating in work takes place throughout most of the adult lifespan (unlike education) and generally follows a consistent schedule (unlike leisure). The relatively weak connection (**β** = 0.10) found between age and cognitive ability in the York model is also notable, as it suggests that variance that would normally be attributed to age may be better explained by cognitive reserve. Thus, building cognitive reserve may potentially offset the process of age-related cognitive decline.

The findings from the current study align with research showing that subcortical regions are a locus for small-vessel disease, which is assumed to have multiple etiologies, including hypertension/hypoperfusion (Hajjar et al., 2011), hypoxia/ischemia (Kang et al., 2022), the buildup of iron (Yan et al., 2013), and demyelination of these highly laminated structures (Li et al., 2023). Previous research has strongly linked volume losses in subcortical regions, including the thalamus, caudate, pallidum, and accumbens, to cognitive decline (Wan et al., 2020). Other studies have revealed memory declines linked to age-related volume loss in the thalamus (Fjell & Walhovd, 2010) and basal ganglia (Hubble, 1998). Smoking has also been found to be associated with greater volume loss in the bilateral thalamus, amygdala, and nucleus accumbens, underscoring the sensitivity of these regions to vascular disruption.

The loading of bilateral subcortical regions in the gray matter disruption factor is consistent with the existing literature on the vascular vulnerability of these structures. Subcortical regions, including the thalamus and basal ganglia, are heavily reliant on small penetrating arteries (Pantoni & Garcia, 1997; Schmidt et al., 2005), making them particularly susceptible to ischemic damage associated with small vessel disease, a primary driver of WMHs (Debette & Markus, 2010; Tuladhar et al., 2016). Given this vascular susceptibility, it follows that subcortical atrophy is linked to cognitive decline, particularly in executive function and processing speed (Li et al., 2024). These findings are consistent with broader frameworks of neural organization, such as the Bilingual Anterior to Posterior and Subcortical Shift model (Grundy et al., 2017), which describes a shift toward greater reliance on posterior and subcortical regions with increased bilingual experience. This may help explain why various studies have suggested that bilingualism acts as a powerful cognitive reserve to help delay the onset of symptoms of dementia (meta-analyses in Anderson et al., 2020; Brini et al., 2020; Paulavicius et al., 2020).

Our results also align with findings based on similar approaches, such as the use of WMHs as the loci for disconnectome maps in older adults. For example, Li et al. (2023) examined diffusion-weighted imaging and WMH lesion masks from 30 healthy subjects from the Human Connectome Project. The authors found that subcortical regions, along with projections to frontal lobes, formed the core of the white matter disconnectome. Moreover, the authors showed that this disconnectome expanded to include motor and parietal cortices with increasing age (Li et al., 2023). Similarly, Rudolph et al. (2024) linked WMH masks to DWI data in 68 cognitively normal adults between 18–78 years of age and found that the disconnectome core focused on the basal ganglia, while the extent of white matter disconnection correlated with declines in fluid cognition. Their results suggested that WMHs were most concentrated around the middle cerebral artery and particularly impacted the anterior thalamic radiations and forceps minor. Thus, there appears to be an emerging consensus that one of the primary impacts of WMHs in aging is to drive disconnection between the subcortical regions and cortex, particularly the frontal lobes.

The data from the current study builds on the link between WMHs and the disconnectome by showing that cognitive decline is mediated by the presence of cognitive reserve, particularly when it is measured with more than one indicator variable. Given that older adults with higher cognitive reserve are less vulnerable to disconnection by lesions than those with lower cognitive reserve, this suggests that these individuals may have more extensive and redundant connections between regions, allowing the superfluous connections to take over when WMHs weaken or destroy primary pathways. In the current study, sex was controlled for in the harmonization process for the combined dataset but not analyzed as a variable of interest.

Previous attempts to examine WMH disconnectomes have relied on datasets with both lesion maps of WMH and white matter tracts derived from diffusion-weighted imaging. Our approach was conceptually similar but took advantage of the LQT toolbox. The LQT approach is optimal in scenarios where the presence of a large lesion, particularly a stroke, would impact the validity of diffusion-weighted scans and make acquiring tracts implausible. For example, diffusion-weighted imaging (DWI), while becoming more accessible, is not universally used across datasets making harmonization of white matter across datasets difficult. Alternatively, through analyzing FLAIR and T1 sequences, the present approach allows for clear inferences to be drawn regarding the influence of extracted lesions. Specifically, the implementation of the LQT allows us to complete pseudo tractography in datasets without diffusion data and was, therefore, the chosen methodology for the current work.

Our data, which was acquired from healthy older adults, suggests that the LQT approach is also suitable for application with WMH lesions; thus, it may not always be necessary to acquire DWI data to compute the disconnectome. Moreover, relying on the LQT method allowed for leveraging and harmonization of datasets that only included structural (T1-weighted) MRIs, thus increasing generalizability and power.

Another strength of our approach was the use of multivariate PLS-PM. This approach offers two notable advantages. First, PLS is an ideal tool for simultaneously modeling multiple, often correlated effects, and it handles multicollinearity well and allows the researcher to correct for multiple comparisons at the level of the latent variable rather than the variable level (Westlund et al., 2008). Second, the path - modeling approach embraced by PLS-PM allows researchers to draw stronger causal interpretations from their data. Based on our findings, we argue that WMHs cause cognitive decline, while cognitive reserve causes higher cognitive ability.

### 4.2 Limitations

While some paths were reliable across datasets (e.g., the relationship between age and gray matter disruption), other paths varied by dataset. For example the age → cognition path was statistically reliable in the ADNI sample, with older participants having lower cognitive ability; however, in all other cases it was not statistically reliable. It is likely that the reliable relationship was a result of an inadvertent selection bias for abnormally healthy individuals who met the strict MRI selection criteria (e.g., no chemotherapy, no heart issues, no chronic pain when lying on one’s back, no neurological issues, etc.). Ultimately, it is possible that this selection process led to the exclusion of people experiencing, for example, atherosclerosis, angina, late-life depression, etc. This may have weakened the ability of the university-led studies, such as York and Cornell, to detect cognitive reserve effects since most of the sample with risk factors for cognitive decline were already excluded. Thus, the impact of cognitive reserve on cognition may be underestimated as a result of the available sample selection. Future studies should include a more representative sample, for example, with less stringent inclusion criteria, and use complementary imaging techniques or low-field MRI. The discrepancies in the models for the different datasets warrant further examination of the relationship between cognitive performance, sex, age, cognitive reserve, and gray matter disruption from WMHs (or disruptions to brain connectivity more broadly). The impact of age on gray matter disruption and cognitive performance does not seem to correlate with the variation in the participants’ ages across different datasets, indicating that age alone may not account for the differences observed. Future research should seek to identify similar measures of cognitive ability and cognitive reserve to test whether these demographic variables led to differences in WMH outcomes between datasets.

While the MMSE is not ideal as a cognitive performance outcome measure, as performance can be at ceiling in healthy individuals, it is reliably reported across many datasets, offering a starting point for harmonization efforts. As more researchers begin to include the Montreal Cognitive Assessment (MoCA; Nasreddine et al., 2005), which is more challenging, it would be useful to extend our approach using datasets that include this measure. Traditionally, education has been a prevalent indicator of cognitive reserve. However, as undergraduate degrees become ubiquitous and graduate degrees more common, this single metric may yield diminishing returns in future studies. Future research should seek to assess additional measures of cognitive reserve to account for developments in educational enrollment.

A notable challenge in our analyses was the limited overlap in behavioral measures across the three datasets. Consequently, in the combined model, cognitive ability and cognitive reserve were each represented by a single indicator, which may have influenced the reliability of several path coefficients. Future research would benefit from a larger sample with a broader range of shared behavioral measures to comprehensively assess cognitive reserve and cognitive decline. Furthermore, a deeper exploration of the specific roles of the different brain regions and functional networks could significantly enhance understanding of how gray matter disruption impacts cognitive function.

### 4.3 Conclusion

The current study is the first to apply the LQT to cognitively normal older adults to assess the impact of WMHs on healthy aging. We found that the LQT’s approach of using regional estimates of disruption to brain connectivity, as an alternative to traditional whole-brain approaches, can be used to predict lower cognitive ability. Furthermore, cognitive reserve predicts higher cognitive ability, especially when captured by a robust measure (CRIq), suggesting a protective effect of building cognitive reserve earlier in life. Integrating information on localized disruption to connectivity and the compensatory effects of cognitive reserve will strengthen future research on WMHs and cognitive decline.

## Data Availability

All data produced in the present study and code for the statistical analyses are available at the research project's OSF page: https://osf.io/px7dy/

https://osf.io/yhzxe/

https://openneuro.org/datasets/ds003592/versions/1.0.13

## Acknowledgements

This research was supported by a Canada Research Chair (Tier II, CRC-2020-00174) and NSERC Discovery Grant (DGECR-2022-00309) to JAEA. Data collection and sharing for this project was funded by the Alzheimer’s Disease Neuroimaging Initiative (ADNI) (National Institutes of Health Grant U01 AG024904) and DOD ADNI (Department of Defense award number W81XWH-12-2-0012). ADNI is funded by the National Institute on Aging, the National Institute of Biomedical Imaging and Bioengineering, and through generous contributions from the following: AbbVie, Alzheimer’s Association; Alzheimer’s Drug Discovery Foundation; Araclon Biotech; BioClinica, Inc.; Biogen; Bristol-Myers Squibb Company; CereSpir, Inc.; Cogstate; Eisai Inc.; Elan Pharmaceuticals, Inc.; Eli Lilly and Company; EuroImmun; F. Hoffmann-La Roche Ltd and its affiliated company Genentech, Inc.; Fujirebio; GE Healthcare; IXICO Ltd.; Janssen Alzheimer Immunotherapy Research & Development, LLC.; Johnson & Johnson Pharmaceutical Research & Development LLC.; Lumosity; Lundbeck; Merck & Co., Inc.; Meso Scale Diagnostics, LLC.; NeuroRx Research; Neurotrack Technologies; Novartis Pharmaceuticals Corporation; Pfizer Inc.; Piramal Imaging; Servier; Takeda Pharmaceutical Company; and Transition Therapeutics. The Canadian Institutes of Health Research is providing funds to support ADNI clinical sites in Canada. Private sector contributions are facilitated by the Foundation for the National Institutes of Health (www.fnih.org). The grantee organization is the Northern California Institute for Research and Education, and the study is coordinated by the Alzheimer’s Therapeutic Research Institute at the University of Southern California. ADNI data are disseminated by the Laboratory for Neuro Imaging at the University of Southern California.

