## Supplemental Material for "Cognitive Reserve Disrupts Cognitive Decline from White Matter Hyperintensities"

Supplemental Materials

### Additional PLS-PM Results

**Table S1**

PLS Regression Results for Cognitive Ability by Dataset

| Dataset | $R^2$ | $\boldsymbol{\beta}_{Age}$ | $\boldsymbol{\beta}_{CR}$ | $\boldsymbol{\beta}_{GMD}$ |
|---|---|---|---|---|
| Combined | 0.05 | 0.06 | 0.06 | -0.23 |
| ADNI | 0.08 | 0.09 | 0.05 | -0.31 |
| York | 0.05 | -0.19 | 0.06 | -0.04 |
| Cornell | 0.05 | 0.12 | 0.20 | -0.27 |

*Note.* Regression results for predicting cognitive ability from age, cognitive reserve, and gray matter disruption. These are the same variables that were used for PLS-PM, and the regression results were generated simultaneously to the PLS-PM results using the plspm package (Sanchez, 2013) in R.

**Table S2**

PLS-PM Unidimensionality Indicators for Gray Matter Disruption by Dataset

| Dataset | Cronbach's alpha | Dillon-Goldstein's rho | 1st Eigenvalue | 2nd Eigenvalue |
|---|---|---|---|---|
| Combined | 0.95 | 0.96 | 23.35 | 11.03 |
| ADNI | 0.85 | 0.87 | 6.63 | 2.85 |
| York | 0.74 | 0.80 | 4.62 | 2.33 |
| Cornell | 0.69 | 0.75 | 4.43 | 2.82 |

**PLS-PM Models**

**Figure S1A**

PLS-PM Model for the ADNI Dataset

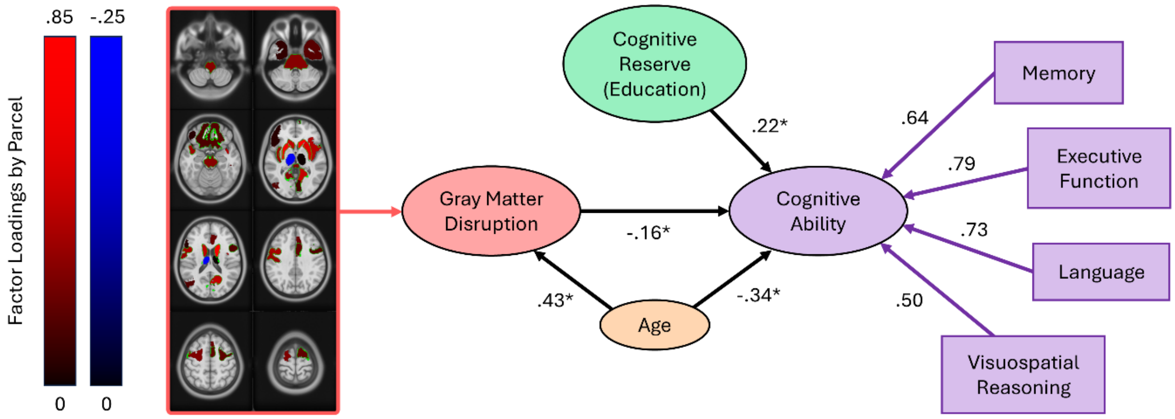


*Note.* This PLS-PM model is based on the ADNI dataset only (*N* = 120). The black arrows represent the directional causal paths between variables specified a priori from past literature for the path model. The coefficients beside the arrows represent the strength of the causal links determined empirically from the data using PLS-PM. Asterisks next to coefficients indicate statistically reliable paths (i.e., the bootstrapped 95% confidence intervals for the coefficients did not include zero). Gray matter disruption indicates the extent to which communication to gray matter regions in the brain is affected by WMHs in white matter tracts connecting to them. The loadings of disruption scores for individual gray matter regions onto the overall gray matter disruption factor are shown on the left. Positive loadings are shown in red, and negative loadings are shown in blue; brighter shades represent greater absolute values.

**Figure S1B**

PLS-PM Model for the Cornell Dataset

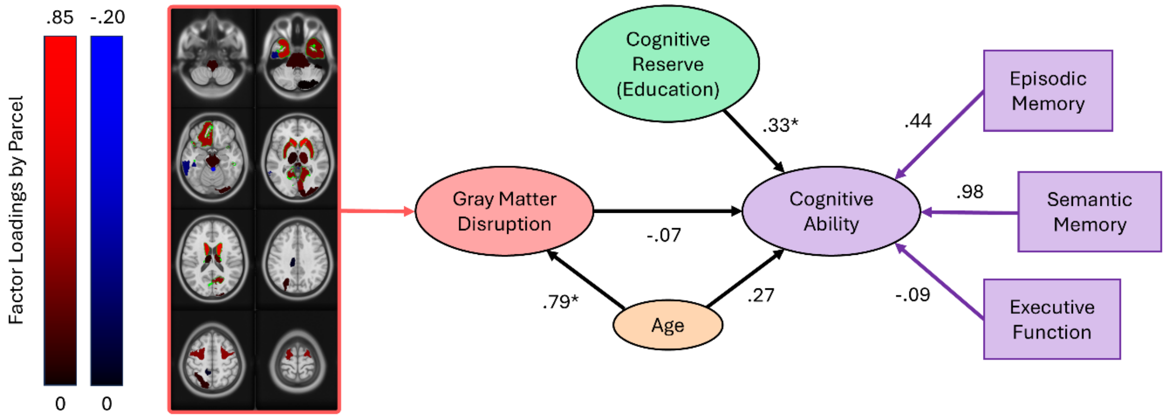


*Note.* This PLS-PM model is based on the Cornell dataset only ($N = 51$). The black arrows represent the directional causal paths between variables specified a priori from past literature for the path model. The coefficients beside the arrows represent the strength of the causal links determined empirically from the data using PLS-PM. Asterisks next to coefficients indicate statistically reliable paths (i.e., the bootstrapped 95% confidence intervals for the coefficients did not include zero). Gray matter disruption indicates the extent to which communication to gray matter regions in the brain is affected by WMHs in white matter tracts connecting to them. The loadings of disruption scores for individual gray matter regions onto the overall gray matter disruption factor are shown on the left. Positive loadings are shown in red, and negative loadings are shown in blue; brighter shades represent greater absolute values.

**Figure S2A**

PLS-PM Model for the ADNI Subset of the Combined Dataset

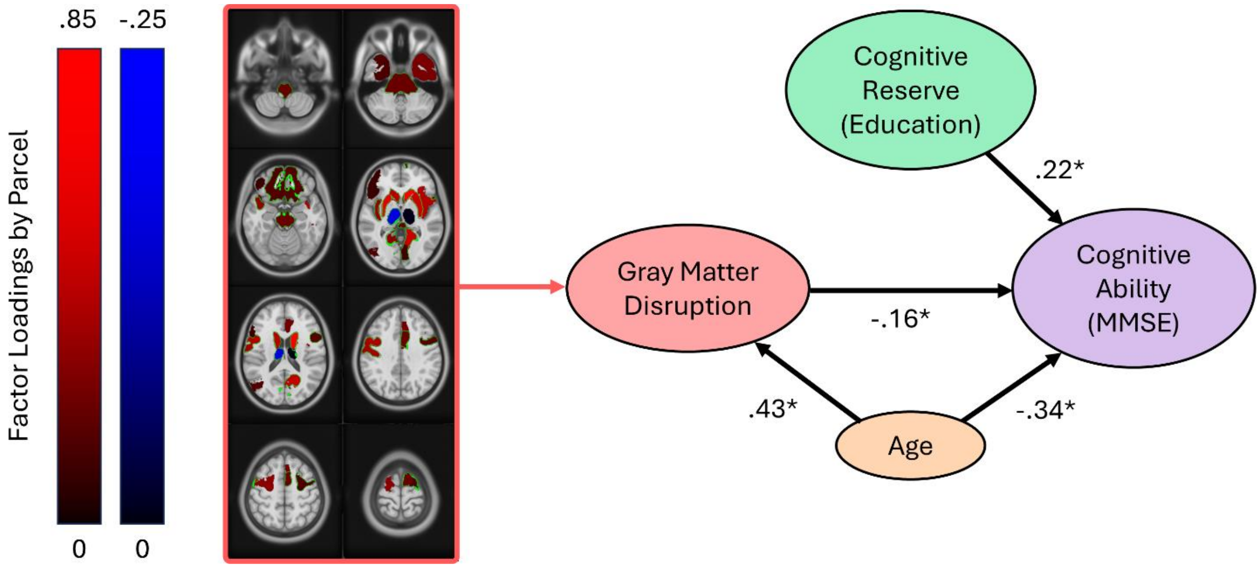


*Note.* This PLS-PM model is based on the data from the Combined model, but only the subset from ADNI participants ($N = 120$). This model therefore differs from the main ADNI PLS-PM model (see Figure S1A) because it uses the measures from the Combined model and not the wider range of measures of cognitive ability used in the main ADNI model. The black arrows represent the directional causal paths between variables specified a priori from past literature for the path model. The coefficients beside the arrows represent the strength of the causal links determined empirically from the data using PLS-PM. Asterisks next to coefficients indicate statistically reliable paths (i.e., the bootstrapped 95% confidence intervals for the coefficients did not include zero). Gray matter disruption indicates the extent to which communication to gray matter regions in the brain is affected by WMHs in white matter tracts connecting to them. The loadings of disruption scores for individual gray matter regions onto the overall gray matter disruption factor are shown on the left. Positive loadings are shown in red, and negative loadings are shown in blue; brighter shades represent greater absolute values.

**Figure S2B**

PLS-PM Model for the York Subset of the Combined Dataset

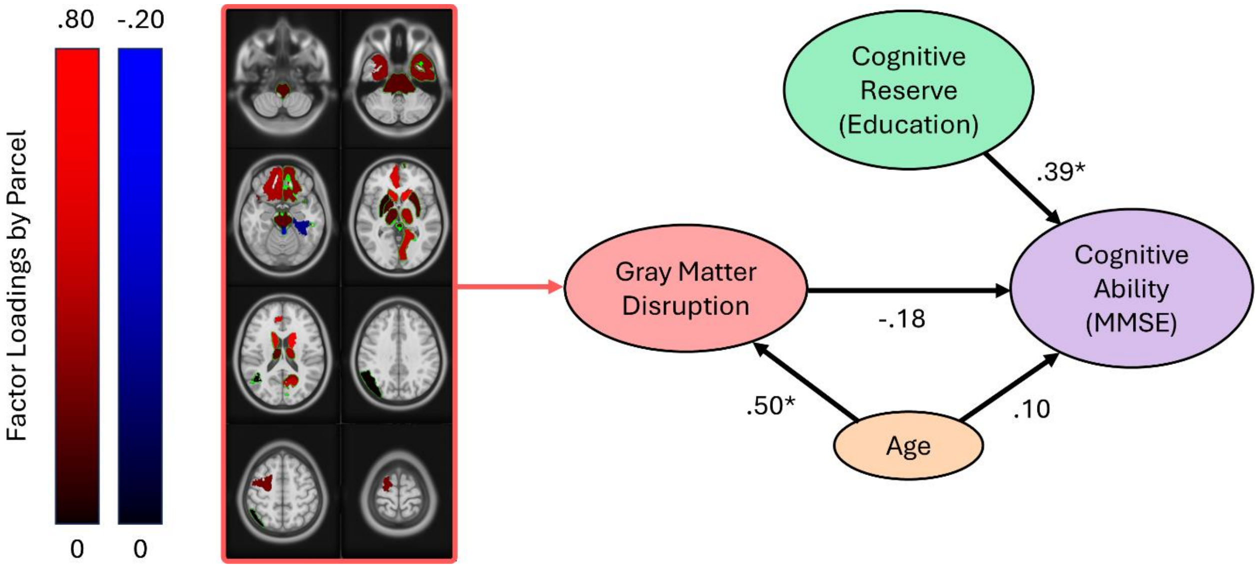


*Note.* This PLS-PM model is based on the data from the Combined model, but only the subset from York participants (*N* = 88). This model therefore differs from the main York PLS-PM model (see Figure 3B) because it uses the measures from the Combined model and not the wider range of measures of cognitive ability and cognitive reserve used in the main ADNI model. The black arrows represent the directional causal paths between variables specified a priori from past literature for the path model. The coefficients beside the arrows represent the strength of the causal links determined empirically from the data using PLS-PM. Asterisks next to coefficients indicate statistically reliable paths (i.e., the bootstrapped 95% confidence intervals for the coefficients did not include zero). Gray matter disruption indicates the extent to which communication to gray matter regions in the brain is affected by WMHs in white matter tracts connecting to them. The loadings of disruption scores for individual gray matter regions onto the overall gray matter disruption factor are shown on the left. Positive loadings are shown in red, and negative loadings are shown in blue; brighter shades represent greater absolute values.

**Figure S2C**

PLS-PM Model for the Cornell Subset of the Combined Model

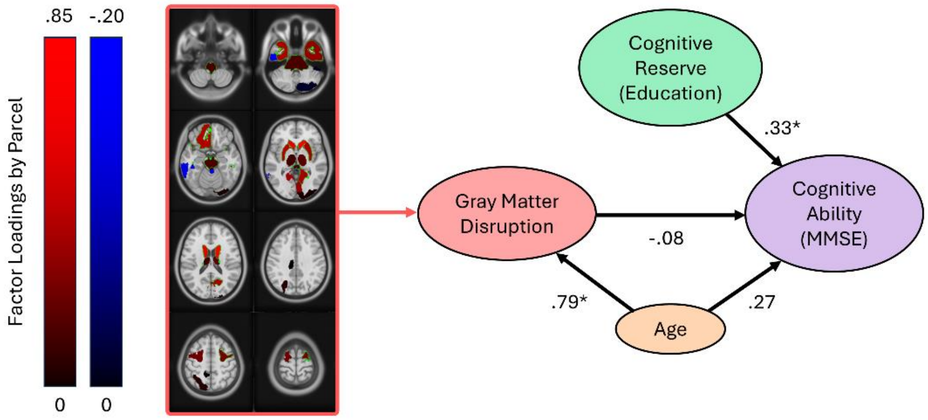


*Note.* This PLS-PM model is based on the data from the Combined model, but only the subset from Cornell participants (*N* = 51). This model therefore differs from the main ADNI PLS-PM model (see Figure S1A) because it uses the measures from the Combined model and not the wider range of measures of cognitive ability used in the main ADNI model. The black arrows represent the directional causal paths between variables specified a priori from past literature for the path model. The coefficients beside the arrows represent the strength of the causal links determined empirically from the data using PLS-PM. Asterisks next to coefficients indicate statistically reliable paths (i.e., the bootstrapped 95% confidence intervals for the coefficients did not include zero). Gray matter disruption indicates the extent to which communication to gray matter regions in the brain is affected by WMHs in white matter tracts connecting to them. The loadings of disruption scores for individual gray matter regions onto the overall gray matter disruption factor are shown on the left. Positive loadings are shown in red, and negative loadings are shown in blue; brighter shades represent greater absolute values.

### Selection Process for Participants from the ADNI Database

Data was screened and downloaded from the ADNI website from 27-30 October 2024. They were 1) screened within the ADNI online database to create a reduced selection of potentially eligible participants, and 2) further screened after the reduced selection was downloaded.

**1. Screening within the ADNI database**

Database entries were filtered using the following search criteria:

- Dataset = ADNI
- Phase = ADNI 3
- Study visit = ADNI3 Year 1 Visit
- Modality = MRI

Entries were then downloaded if they had one of the following descriptions:

- T1 scan descriptions:
    - 3D T1 Sag
    - Accelerated Sagittal MPRAGE
        - For 010_S_6748, misspelled as “Satittal”
    - Accelerated Sagittal MPRAGE L>>R
    - Accelerated Sagittal MPRAGE Phase A-P
    - Accelerated Sagittal MRPAGE REPEAT
    - Accelerated Sagittal MPRAGE MPR Cor
    - Accelerated Sagittal MPRAGE MPR Tra
    - Accelerated Sagittal MPRAGE ND
    - REPEAT Accelerated Sagittal MPRAGE
    - Sagittal 3D Accelerated 0 angle MPRAGE
    - Sagittal 3D Accelerated MPRAGE

    - Accelerated Sag IR-FSPGR
    - Accelerated Sagittal IR-FSPGR
    - ORIG Accelerated Sag IR-FSPGR
    - Sag Accel IR-FSPGR
- FLAIR scan descriptions:
    - Axial T2-FLAIR
    - FLAIR
        - Note: this one was recorded in the axial plane (I believe this just means the superior/inferior dimension is the one with reduced resolution, whereas the rest are sagittal so it would be the left/right dimension)
    - ORIG Sagittal 3D FLAIR
    - Sagittal 3D 0 angle FLAIR
    - Sagittal 3D FLAIR
        - For 010_S_6748, misspelled as “Saggital”
    - Sagittal 3D FLAIR phase A-P
    - Sagittal 3D FLAIR MPR Cor
    - Sagittal 3D FLAIR MPR Tra

**2. Screening the downloaded files**

Firstly, participants lacking either a T1 scan or a FLAIR scan were removed.

Next, participants with multiple T1 scans and/or multiple FLAIR scans were identified. Many participants had multiple scans with same description. In addition, some participants had multiple scans of one type but with somewhat different descriptions; these were the specific participants:

- Had both Accelerated_Sagittal_MPRAGE and Accelerated_Sagittal_MPRAGE_L__R on the same date:
    - 007_S_2394

- Had both Accelerated_Sagittal_MPRAGE and Accelerated_Sagittal_MPRAGE_ND on the same date:
  - 013_S_4580, 013_S_6206
  - 014_S_2308, 014_S_6076, 014_S_6145, 014_S_6199, 014_S_6522, 014_S_6765
  - 022_S_5004, 022_S_6069, 022_S_6716, 022_S_6796
  - 070_S_6229, 070_S_6236, 070_S_6911
  - 082_S_2121, 082_S_4224, 082_S_4428, 082_S_5282, 082_S_6563
  - 100_S_6039
  - 116_S_4199, 116_S_4855, 116_S_6100, 116_S_6133, 116_S_6428, 116_S_6543, 116_S_6550, 116_S_6775
  - 141_S_1052, 141_S_2333, 141_S_4160, 141_S_6008, 141_S_6061, 141_S_6178, 141_S_6779
- Both  Accelerated_Sag_IR-FSPGR and Sagittal_3D_FLAIR had extra versions with the same name and "ORIG_" added to the start, and the same dates:
  - 021_S_6896, 021_S_6994
- The FLAIR is Axial_T2-FLAIR, but no other issues:
  - 022_S_6013
- Had both 3D_T1_SAG and Sagittal_3D_Accelerated_MPRAGE, as well as both FLAIR and Sagittal_3D_FLAIR:
  - 123_S_4127
- Had both Accelerated_Sagittal_MPRAGE and Accelerated_Sagittal_MPRAGE_REPEAT
  - 137_S_4536, 137_S_4631
- Has three of each (Both Accelerated_Sagittal_MPRAGE and Sagittal_3D_FLAIR each have additional versions with _MPR_Cor and _MPR_Tra added)
  - 168_S_6874, 168_S_6875

For each participant, only the best T1 scan and the best FLAIR scan were selected for processing. In order to determine which scan was best, the DICOM files were converted to the NIfTI file format and then assessed as follows:

- Removed the one ending in _L__R; it looked like the other one.
- Removed the ones beginning in ORIG_; they looked like the other ones.
- Kept the ones ending in _REPEAT; they looked almost like the others and I assume the second scan was due to an issue with the first one.
- Removed those ending in _MPR_Cor and _MPR_Tra; they looked like the other ones.
- Removed the ones labelled 3D_TA_Sag and FLAIR; they looked like the other ones.
- Deleted 022_S_6013, which had Axial_T2-FLAIR; it was quite blurry.
- For all those with two or more scans of the same name, I checked both (or all) images in Mango. If there was no important difference, I kept the last one (the .nii file and .json file end in an "a" before the file extension), and then removed the "a" from the end of the .nii and .json files names. If one image had obvious defects in the recording, I went with the other one; only two had obvious defects.

Finally, one participant was removed following visual examination because the FLAIR scan was too blurry to be usable.
